# Long Term Humoral Immunity Decline in Hemodialysis Patients Following SARS-CoV-2 Vaccination

**DOI:** 10.1101/2021.12.01.21265957

**Authors:** Eibhlin Goggins, Binu Sharma, Jennie Z. Ma, Jitendra Gautam, Brendan Bowman

**Author notes:** Funding Source: No External Funding.

## Abstract

Dialysis patients are extremely vulnerable to SARS-CoV-2 infection with high rates of hospitalization and mortality rates estimated at 20-30%. In January of 2021, the University of Virginia Dialysis Program initiated a program wide vaccination campaign administering Pfizer BioNTech mRNA SARS-CoV-2 (BNT162b2) vaccine. To characterize the time-dependent decline in humoral immunity, we performed a prospective cohort study measuring serial monthly semi quantitative IgG antibody levels to the SARS-CoV-2 spike protein receptor binding domain in fully vaccinated in-center hemodialysis patients. Measurements were taken beginning at 2 months post full vaccination through 6 months after full vaccination. Early results showed similar seroconversion rates as prior studies with 88% obtaining positive antibody levels. Those with prior infection obtained the highest antibody levels. Over the ensuing months, patient antibody levels declined at an adjusted average rate of 31% per month. At the conclusion of the study, 40% of patients remaining in the cohort possessed either negative or borderline IgG antibody levels. Projecting future antibody levels based on the slopes of antibody level decay suggests 65% of the cohort will progress to borderline or negative antibody levels at 10 months post full vaccination. In summary, we studied long term vaccine response following vaccination with the BNT162b2 mRNA vaccine in hemodialysis patients. Our data adds to the limited pool of data in this patient population and will help to inform the discussion about vaccine booster needs and frequency.

## Introduction

Among patients with end stage kidney disease (ESKD), morbidity and mortality from infection with SARS-CoV-2 is high.^1^ Particularly, dialysis patients are at high risk of infection from SARS-CoV-2 due to unavoidable exposure from frequent encounters with the healthcare system and impaired humoral and cellular immune function. Initial trials of the Pfizer-BioNTech COVID-19 vaccine (BNT162b2) demonstrated robust antibody responses in participants; however, patients with ESKD were excluded from these studies.^2^

Data regarding SARS-CoV-2 vaccine humoral response in dialysis patients has recently been reviewed.^3^ Briefly, 80-90% of patients with ESKD attain detectable IgG antibodies to the spike protein receptor binding domain component of SARS-CoV-2.^4-6^ These rates, while impressive, remain lower than those observed in the general population. Dialysis patients also attain lower antibody levels in response to mRNA vaccines compared to healthy controls.^7^ Despite the majority of dialysis patients attaining a positive antibody response, the strength or durability of humoral immunity following a standard two shot vaccine regimen is not known. Given known underlying immune dysfunction of patients with ESKD, it is likely that long term vaccine strategies in dialysis patients may differ from that of the general public.

In January of 2021, the University of Virginia began a dialysis program-wide vaccination campaign exclusively using BNT162b2 (courtesy of the Virginia Department of Health) achieving an 80% vaccination rate among all prevalent dialysis patients.^8^ From this cohort, we selected a subset of patients to prospectively study serial antibody levels from two to six months following full vaccination. Here, we report results of serial monthly antibody levels, slope of antibody decline and qualitative population loss of detectable humoral antibody response in this selected subset.

## Methods

### Study Population

69 patients undergoing in-center hemodialysis were confirmed as fully vaccinated at the sole University of Virginia study site / dialysis center. Of these, 35 adults (>18 years) were enrolled in this study. Sample size was based on pragmatic considerations of sample volume processing capability. All participants had received two doses of the BNT162b2 vaccine between January and February 2021. Patients dialyzing for acute kidney injury and those with active infection or suspected SARS-CoV-2 infection requiring isolation were excluded at enrollment (Supplemental Figure 1).

### Sample Collection and Assessment

Samples from participants were obtained on a monthly basis beginning at a mean of 9.1 weeks post full vaccination (defined as >14 days following second immunization) on designated collection dates for each dialysis shift (MWF or TTS). A 10 mL EDTA tube was collected from each patient’s dialysis blood line during dialysis treatment, stored in a designated research refrigerator and processed within 8 hours of initial collection. Tubes were centrifuged at 3000 rpm (1620rcf) for 10 minutes in the swing bucket rotor (S4180) at 4°C using a Beckman GS-15R centrifuge. Plasma obtained was stored in −80°C in 0.5 mL aliquots until further analysis.

All monthly EDTA plasma samples were tested for anti SARS-CoV-2 antibodies against anti-spike S1 domain using the commercially available Anti-SARS-CoV-2 QuantiVac ELISA (IgG) from Euroimmun (EUROIMMUN US, Inc., 1 Bloomfield Ave, Mountain Lakes, NJ, USA). Assays were run and results interpreted as per the manufacturer’s guidelines. Samples above detection limits were re-run with further dilution (1:5 or 1:10) in the sample buffer as recommended by the manufacturer. Based on the manufacturer’s recommendation, final test results were presented as the internationally harmonized binding antibody units (BAU/mL).^9^ BAU/mL was obtained by multiplying the Relative Unit (RU/mL) by a factor of 3.2. Final test results were considered negative for BAU/mL (< 25.6), borderline for BAU/mL (≥25.6 and <35.2) and positive for BAU/mL (≥35.2).^9^

To assess for undiagnosed prior infection and confirm reported histories of prior infection, the Bio-Rad Platelia SARS-CoV-2 Total Ab assay (Bio-Rad Laboratories, Inc., Hercules, CA, USA) was used for qualitative detection of total antibodies (IgM/IgG/IgA) to SARS-CoV-2 nucleocapsid protein. Testing was run from EDTA plasma and limited to each patient’s initial sample only. Recombinant SARS nucleocapsid protein is used in the assay to capture total antibodies in a one-step antigen capture format followed by detection.

### Data Collection

Demographic data including age, sex, race/ethnicity, and BMI and clinical data including comorbidities, use of immune suppressive medication, history of malignancy, and history of transplantation were obtained from the Electronic Health Record (Table 1). Clinical information including dialysis vintage was obtained from the dialysis-specific electronic medical record system. Prior COVID-19 infection information was collected from a designated tracking file in the dialysis unit and verified with SARS-CoV-2 nucleocapsid protein assay results.

**Table 1:** Patient characteristics by prior COVID-19 infection and immune suppression

| Characteristics | Prior COVID-19 infection |  | Immune suppression |  | Overall<br>(N=35) |
| --- | --- | --- | --- | --- | --- |
|  | No<br>(N=29) | Yes<br>(N=6) | No<br>(N=26) | Yes<br>(N=9) |  |
| Age | 62.55 ± 11.20 | 59.33 ± 11.00 | 64.54 ± 9.360 | 54.67 ± 12.87 | 62.00 ± 11.07 |
| Female | 15 (51.7%) | 3 (50.0%) | 13 (50.0%) | 5 (55.6%) | 18 (51.4%) |
| Race |  |  |  |  |  |
| African American | 16 (55.2%) | 5 (83.3%) | 17 (65.4%) | 4 (44.4%) | 21 (60.0%) |
| White | 13 (44.8%) | 0 (0%) | 8 (30.8%) | 5 (55.6%) | 13 (37.1%) |
| Asian | 0 (0%) | 1 (16.7%) | 1 (3.8%) | 0 (0%) | 1 (2.9%) |
| Etiology ESRD |  |  |  |  |  |
| DM | 9 (31.0%) | 1 (16.7%) | 8 (30.8%) | 2 (22.2%) | 10 (28.6%) |
| GN | 4 (13.8%) | 0 (0%) | 2 (7.7%) | 2 (22.2%) | 4 (11.4%) |
| HTN | 5 (17.2%) | 2 (33.3%) | 5 (19.2%) | 2 (22.2%) | 7 (20.0%) |
| Other | 11 (37.9%) | 3 (50.0%) | 11 (42.3%) | 3 (33.3%) | 14 (40.0%) |
| Dialysis Vintage (Yr)<br>Median [Q1-Q3] | 4.48 [1.64, 8.90] | 4.18 [3.04, 10.17] | 4.78 [2.06, 8.33] | 4.20 [0.79, 10.83] | 4.48 [1.84, 9.87] |
| Access Type |  |  |  |  |  |
| AVF | 16 (55.2%) | 2 (33.3%) | 14 (53.8%) | 4 (44.4%) | 18 (51.4%) |
| CVC | 13 (44.8%) | 4 (66.7%) | 12 (46.2%) | 5 (55.6%) | 17 (48.6%) |
| Solid Organ Transplants | 5 (17.2%) | 1 (16.7%) | 2 (6.16) | 4 (45.4%) | 6(17.1%) |
| Cancer History | 8(72.4%) | 4(66.7%) | 5(19.2%) | 5(55.6%) | 10(28.6%) |
| Comorbidities |  |  |  |  |  |
| DM | 17 (58.6%) | 4 (66.7%) | 16 (61.5%) | 5 (55.6%) | 21 (60.0%) |
| HTN | 29 (100%) | 6 (100%) | 26 (100%) | 9 (100%) | 35 (100%) |
| CVA | 8 (27.6%) | 0 (0%) | 6 (23.1%) | 2 (22.2%) | 8 (22.9%) |
| COPD | 4 (13.8%) | 0 (0%) | 4 (15.4%) | 0 (0%) | 4 (11.4%) |
| CHF | 13 (44.8%) | 4 (66.7%) | 12 (46.2%) | 5 (55.6%) | 17 (48.6%) |
| MI | 6 (20.7%) | 2 (33.3%) | 6 (23.1%) | 2 (22.2%) | 8 (22.9%) |
| PAD | 3 (10.3%) | 0 (0%) | 1 (3.8%) | 2 (22.2%) | 3 (8.6%) |
| Obesity(BMI>30) | 12 (41.4%) | 2 (33.3%) | 11(42.3%) | 3(33.3%) | 14(42.86%) |
| Immune suppression | 8 (27.6%) | 1 (16.7%) | — | — | 9 (25.7%) |
| Prior COVID-19 infection | — | — | 5 (19.2%) | 1 (11.1%) | 6 (17.1%) |
| BAU/ml (Month 2) | 371.0 ± 407.6 | 1892 ± 866.3 | 741.0 ± 827.2 | 398.6 ± 604.4 | 647.6 ± 779.2 |
| BAU/ml (Month 6) | 66.99 ± 66.79 | 710.4 ± 450.1 | 222.6 ± 348.9 | 60.71 ± 75.3 | 177.9 ± 306.2 |

### Statistical Methods

Data were summarized as mean and standard deviation or median (25^th^, 75^th^ percentiles) for continuous variables and as frequency and percentage for categorical variables. The main objective was to estimate the slope of antibody level decline from the time of full immunization to 6 months post full immunization. To analyze the change in antibody levels over time, a linear mixed model with random slope and random intercept was used for longitudinal antibody levels to account for patient-specific changes and variation over the entire follow-up period. The antibody level was natural log transformed for its skewed distribution before the analysis. Univariate models were used to test the association of the trajectories of antibody levels with patient characteristics including prior COVID-19 infection, immune suppression, gender, age, race, Charlson Comorbidity Index (CCI), and access type. The interactive effect of prior COVID-19 infection and immune suppression variable with time were also tested. A multivariable model was used to estimate the slope of antibody levels by adjusting for selected patient characteristics, including age, gender, prior COVID-19 infection and immune suppression because of the small sample sizes. In addition, based on the estimated intercepts and slopes for each subject from the unadjusted model, 10-month antibody level was projected and plotted in a spaghetti graph versus the observed value. p < 0.05 was considered significant. All analyses were performed using software R (version 3.6.3).

## Results

Table 1 provides the clinical characteristics of all study subjects. The mean age was 62.0 years. 51.43% were women and 60% were African American. Mean dialysis vintage was ∼4.5 years. Three participants previously tested positive for COVID-19 and three additional prior infection cases were identified using SARS-CoV-2 nucleocapsid protein assay results, yielding 17% of the study population with prior infection. Nine subjects were defined as immune suppressed at baseline based on current immune suppressive medication use or predisposing medical condition. One patient had a prior infection and was also categorized as immune suppressed. Over the course of the study, one patient withdrew consent, another received a successful transplant, and three patients died from causes unrelated to SARS-CoV-2 infection.

A total of 153 samples were collected from 35 patients. Out of 35, 25 (71%) patients completed all five sample collections. Baseline (i.e., month 2) spike protein IgG levels in BAU/mL are presented in Table 1. The mean baseline antibody level was 647.59 BAU/mL, and 87.88% (29/33) of patients were considered qualitatively positive based on manufacturer provided cutoffs (Figure 2). Two patients were negative at baseline and an additional two had borderline results, yielding an 88% overall initial positive response. Of the initial four borderline or negative subjects, two were categorized as immune suppressed and another had a history of malignancy, consistent with prior studies.

At three, four, five and six months following full vaccination, the average antibody levels fell to 491.4, 365.6, 302.0 and 177.9 BAU/mL, respectively (Figure 1). As expected, the antibody levels in log-scale significantly declined over time (p < 0.05). Further, the unadjusted results (Table 2) show that prior COVID-19 infection was significantly associated with attained antibody level (p < 0.001) but that immune suppression was not (p = 0.12). On average, patients with prior COVID-19 infection had a 9 times higher antibody level than those without. Age, gender, dialysis vintage, Charlson Comorbidity Index, immune suppression and access type were not significantly associated with antibody level. Race was significantly associated; however, the relationship was spurious as 5 of 6 patients with prior infection were African American (Table 1). The interactive effect of prior COVID-19 infection and immune suppression with time were not significant (Table 2), suggesting the log linear decay of antibody levels in these patients were similar.

**Figure 1:**
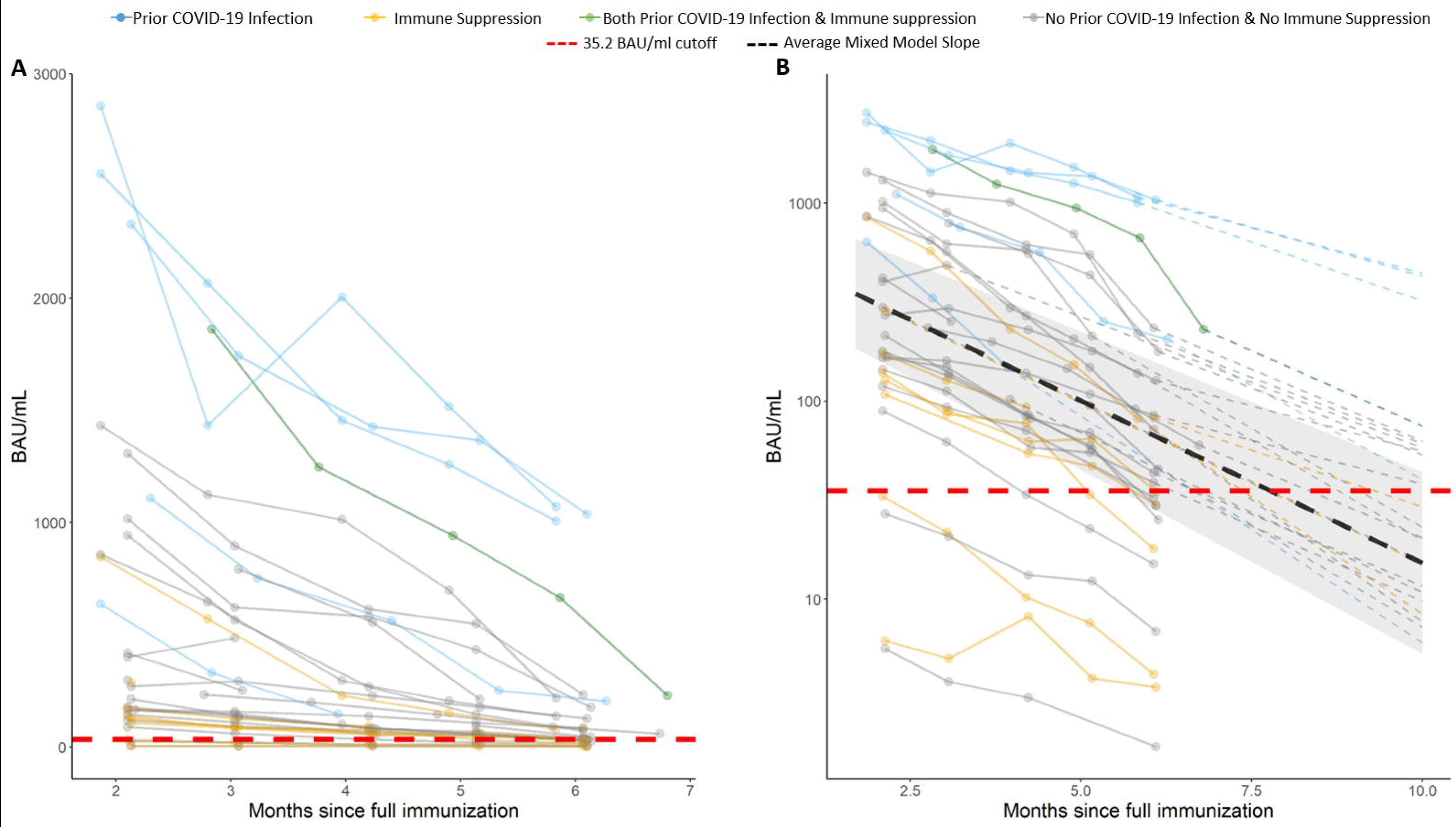
A) Observed antibody level of SARS-CoV-2, the lines are colored by prior COVID-19 infection and immune suppression status. B) Logarithmic scale (y axis) prediction graph on individuals post 10 month since full vaccination. The dark lines are observed values and the dotted lines are predicted values (dashed line not shown for subjects who are “negative” or “borderline” at month 6). Individual intercept and slope estimated from unadjusted linear mixed model were used for prediction. The cutoff for borderline/negative antibody level is 35.2 (red dashed line).

**Table 2:** Univariate and Multivariate results from Linear Mixed Model of log antibody level

| Effects | Univariate Analysis |  | Multivariable Analysis |  |
| --- | --- | --- | --- | --- |
|  | Estimates<br>(95% CI) | p-value | Estimates<br>(95% CI) | p-value |
| Time (per month) | -0.38 (-0.43, -0.32) | <0.001 | -0.37 (-0.43, -0.32) | < 0.001 |
| Age (Per year) | -0.02 (-0.07, 0.2) | 0.283 | -0.03 (-0.08, -0.00) | 0.075 |
| Male | 0.41 (-0.61, 1.44) | 0.423 | 0.27 (-0.51, 1.06) | 0.502 |
| Race |  |  |  |  |
| No African American (reference) |  |  |  |  |
| African American | 1.03 (0.05, 2.01) | 0.044 |  |  |
| Dialysis Vintage |  |  |  |  |
| >= 5 years (reference) |  |  |  |  |
| < 5years | -0.43 (-1.43, 0.56) | 0.398 |  |  |
| Charlson Comorbidity Index (Per unit) | -0.03 (-0.23, 0.17) | 0.773 |  |  |
| Access |  |  |  |  |
| AVF (reference) |  |  |  |  |
| CVC | -0.23 (-1.2, 0.80) | 0.651 |  |  |
| Immune suppression (Yes) | -0.90 (-2.01, 0.20) | 0.120 | -1.11 (-2.07, -0.14) | 0.038 |
| Prior COVID-19 infection (Yes) | 2.19 (1.09, 3.31) | <0.001 | 1.97 (0.95, 3.00) | 0.001 |
| Time * Prior COVID-19 infection | 0.04 (-0.09, 0.17) | 0.581 |  |  |
| Time * Immune Suppression | -0.00 (-0.12, 0.11) | 0.959 |  |  |

Adjusted multivariable linear mixed model included time, age, gender, prior COVID-19 infection and immune suppression. Race was not included as noted above. After adjustment, time, prior COVID-19 infection, and immune suppression were significantly associated (p < 0.05) and age was marginally associated (p = 0.075) with the trajectory of antibody level (Table 2). Keeping all other variables constant, the antibody level per month decayed by an average of 31%. Older patients experienced greater decay in the antibody levels, at an additional 4% decline for one-year increment in age. Immune suppressed patients, on average, had a 65% lower antibody level compared to patients without immune suppression and patients with prior COVID-19 infection had 5 times higher antibody levels than infection naïve patients (Table 2).

Based on manufacturer’s antibody level cutoffs at six months post full immunization (positive, borderline, negative), 61% (17/28) of patients maintained positive antibody levels while 39% (11/28) had borderline or negative antibody levels (Figure 2). Additionally, the prediction of antibody level at month 10 post full vaccination demonstrated that more than 65% of the study population is anticipated to progress to borderline or negative antibody status (Figure 1).

**Figure 2:**
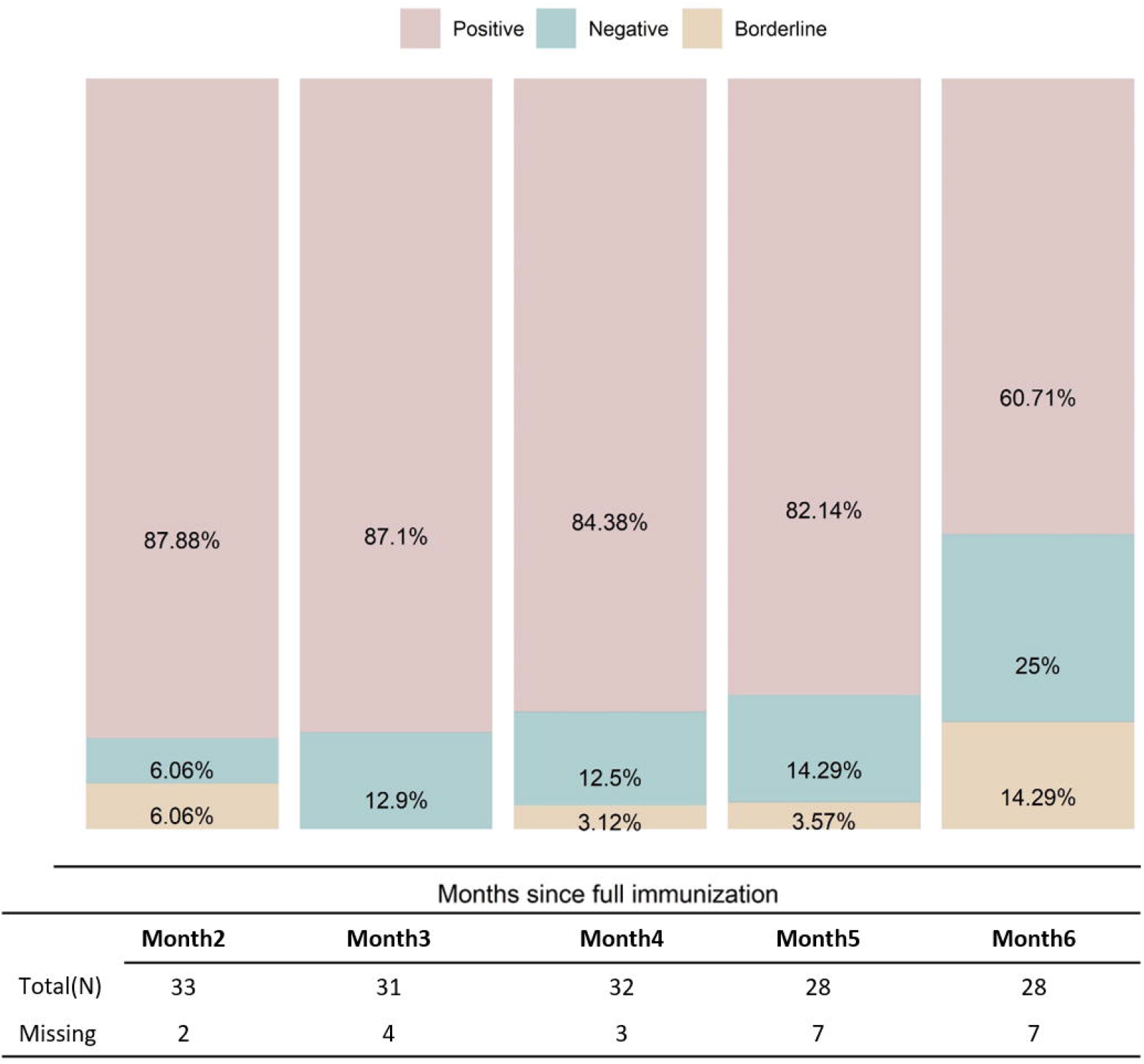
Percentage of persons who were positive, negative, and borderline based on antibody level by time-point from baseline sample collection of month ∼2 post full vaccination to month ∼6 post full vaccination. Percentages are calculated based on total (N) of the table.

## Discussion

Data on COVID-19 vaccine response and, importantly, durability in dialysis patients remains scarce. Thus, the goal of our study was to evaluate long term SARS-CoV-2 spike protein antibody response decay curves in a cohort of early vaccinated prevalent dialysis patients. We analyzed the trajectory of long term IgG spike protein antibody decline and explored the association of antibody level with patient characteristics. Our data also confirms previously described findings showing lower rates of seroconversion in dialysis patients as well as antibody level attenuation associated with immune suppression and advancing age.

We observed a generally stable decline in IgG spike protein antibody levels from month to month regardless of subgroup or initial antibody peak. This relatively stable decay rate suggests that peak attained antibody level post vaccination is a predictive factor determining the duration of detectable IgG spike protein antibody levels. In fact, none of the previously infected patients are projected to lose detectable antibody levels at 10 months post full immunization (Figure 1B). As shown in earlier studies, subjects with prior COVID-19 infection developed high antibody levels early on (Figure 1) and these remained higher 6 months post full vaccination in prior infected patients relative to infection naïve patients (888.01 BAU/mL versus 61.01 BAU/mL respectively) (Table 1). Similarly, the ROMANOV study found HD patients with a prior history of COVID-19 had a vaccine response identical to their healthy controls, but response was significantly lower in their infection naïve HD patients.^10^ In fact, the vast majority of the study cohort, approximately 65%, are likely to experience loss of detectable IgG spike protein antibody at 10 months post full vaccination. Our real world data through six months of full vaccination already demonstrates nearly 40% of our cohort have antibody levels at borderline or negative thresholds. While we anticipated immune suppressed patients to lose detectable antibody levels, the 40% of patients already in the borderline or negative group are mostly drawn from the infection naïve group, not the immune suppressed group.

Immunity and vaccine effectiveness are determined by many factors, not solely humoral components. However, there appears to be an inversely proportional relationship between antibody levels and symptomatic SARS-CoV-2 infection.^11^ Therefore, dialysis patients, highly vulnerable to SARS-CoV-2, may benefit from planned boosters at the population level. Data on response to boosters in dialysis patients is limited, but a cohort of French dialysis patients demonstrates significant increases in spike protein IgG levels following a third dose of BNT162b2 given at a median of 50 days following a protocol second shot of BNT162b2.^12^ Increased proportional response to the third shot was seen in those with lower initial levels and longer duration of time to the third shot. How a third shot may affect antibody response and durability in dialysis patients 6 months or more after full immunization is not known.

Our study has limitations which may limit generalizability. Notably, we had a small sample size and a non-representative sample. Our population was majority African American and had a large proportion of patients considered immune suppressed. Because we obtained early access to vaccine relative to the U.S. dialysis population, we were not able to obtain pre-vaccination or peak antibody levels at the typical 14 days after second immunization. We also reported solely on antibody response to BNT162b2 and not other COVID-19 vaccines. Lastly, although data has described a correlation between spike protein IgG levels and infection vulnerability, protective antibody levels have not been clearly determined. Thus, results should be cautiously interpreted. Strengths of our study include the long term nature, diverse clinical background of our cohort, and the relatively complete data set allowing the development of antibody level trajectory curves.

In conclusion, we present long term IgG spike protein antibody decline rates in response to vaccination with BNT162b2. These results suggest that dialysis patients vaccinated with BNT162b2 and without prior infection may benefit from receipt of a booster dose.

## Supporting information

COI_Disclosure_BB

COI_Disclosure_BS

COI_Disclosure_JG

COI_Disclosure_JM

COI_Disclosure_EG

STROBE

## Data Availability

Data is not freely available externally. Please contact author if interested in de-identified data set for research purposes

## Acknowledgements & Funding

This study was funded by the UVA Division of Nephrology Clinical Research Center. The authors wish to gratefully acknowledge the support of several individuals at UVA that contributed to this study. First, Elizabeth Kirkland for providing IRB and study support as lead clinical research coordinator. Danielle Wentworth MSN, FNP for assistance consenting and obtaining samples. The UVA dialysis center managers and staff who assisted in obtaining monthly samples. Drs. Mark Okusa and Julia Scialla for providing study design feedback and support. And most importantly, our patients who agreed to join our study.

---

**Supplemental Figure 1:**
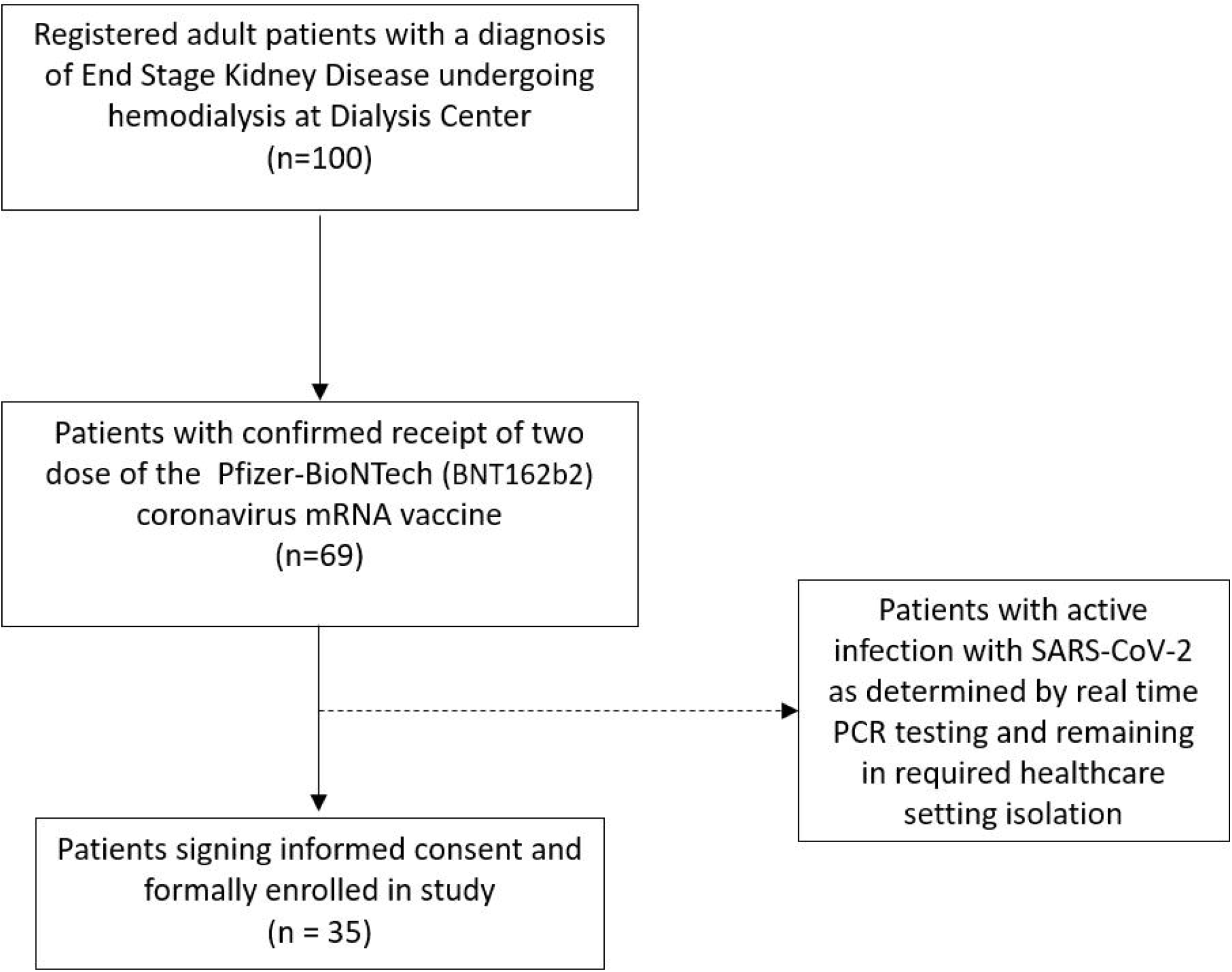
Study Participant Inclusion Flow Chart

**Supplemental Figure 2:**
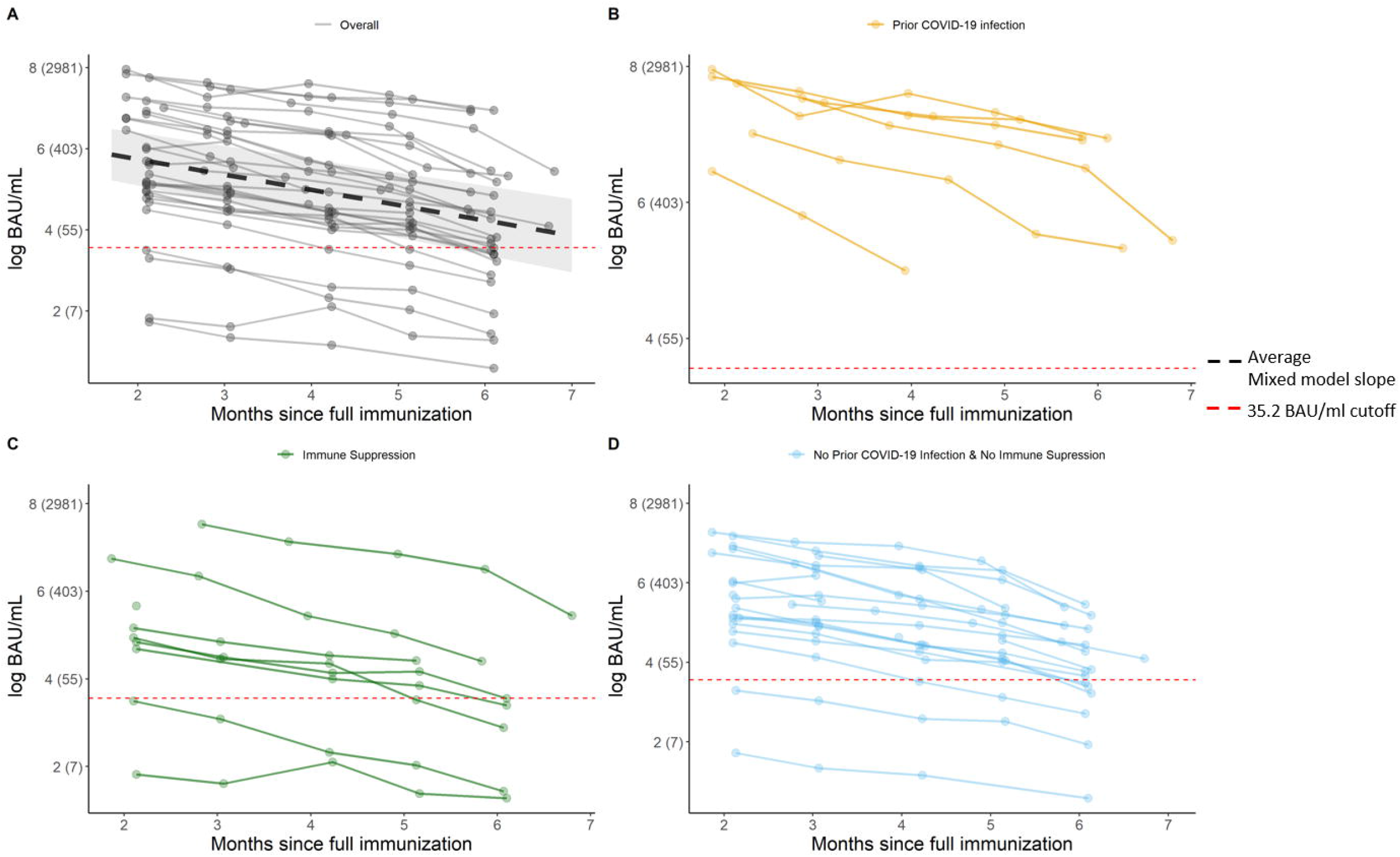
Spaghetti plot for observed antibody levels of SARS-CoV-2 over time from month ∼2 to month ∼6 post full immunization (i.e., 14 days after second dose of vaccination) in (A) Overall sample, (B) Prior COVID-19 infection group, (C) Immune suppression group, (D) Naïve group, No prior infection, and No immune suppression group. Antibody levels are presented in log scale and actual values are shown inside parentheses. The cutoff for borderline/negative antibody level was defined according to manufacturer i.e. 35.2 BAU/mL (red dashed line). The estimated unadjusted slope by linear mixed model including all 35 subjects is shown in A (thick black dashed line).

